# Maternal transfer of IgA and IgG SARS-CoV-2 specific antibodies transplacentally and via breastfeeding

**DOI:** 10.1101/2021.12.21.21267733

**Authors:** Mohammad M. Sajadi, Narjes Shokatpour, Allison Bathula, Zahra Rikhtegaran Tehrani, Allison Lankford, Madeleine Purcell, James D. Campbell, Elizabeth Adrianne Hammershaimb, Kristopher B. Deatrick, Casey Bor, Dawn M. Parsell, Colleen Dugan, Andrea R. Levine, Sabrina C. Ramelli, Daniel S. Chertow, Daniel L Herr, George K. Lewis, Alison Grazioli

## Abstract

Although there have been many studies on antibody responses to SARS-CoV-2 in breastmilk, very few have looked at the fate of these in the baby. We carried out a study in 22 mother/baby pairs (mothers who breastfed and who were SARS-CoV-2 vaccinated before or after delivery) looking at mother blood, mother milk, baby blood, baby nose, and baby stool. Breastfed infants only acquired systemic anti-SARS-CoV-2 IgG antibodies if their mothers were vaccinated antepartum. None of the infants had SARS-CoV-2-specific IgA in the blood, but surprisingly, half of the infants in the Antepartum group had high titer SARS-CoV-2-specific IgA in the nose that exceeded titers found in breastmilk. Vaccination antepartum followed by breastfeeding appears to be the best way to provide systemic and local anti-SARS-CoV-2 antibodies for infants.

## Introduction

SARS-CoV-2 vaccines are available to those over the age of 5; however, infants can theoretically be protected with maternal antibodies. Although COVID-19 vaccination is recommended for pregnant women,^1^ some may choose vaccination postpartum or not at all. Given the ability of maternal antibodies to cross into the fetus antepartum, and for breastmilk to contain antibodies, we decided to determine the distribution and fate of such antibodies in mothers and their infants.

## Methods

Mother/infant pairs at the University of Maryland Medical Center were recruited: mothers vaccinated antepartum who breastfed (Antepartum group), and mothers who breastfed but vaccinated postpartum (Postpartum group). IgG and IgA SARS-CoV-2 antibody levels in maternal blood and milk, as well as infant blood, nose, and stool were determined by ELISA.^2,3^

All mothers provided written informed consent; the study was approved by the University of Maryland institutional review board. Statistical analysis was performed with GraphPad Prism 5. Antibody titers between groups were tested using the 2-tailed Mann-Whitney test, with P<.05 considered significant.

## Results

Ten mother/infant pairs (Antepartum group) and 12 pairs (Postpartum group) had sampling at one time point (Table 1). SARS-CoV-2 specific IgG and IgA were detected in the mothers’ blood and milk demonstrating successful vaccination (Figure 1A and 1B). Breastfed infants in the Antepartum group had SARS-CoV-2-specific IgG in their blood and noses (Figure 1A and 1C). Breastfed infants in the Postpartum group had no detectable SARS-CoV-2-specific IgG in their blood or noses (Figure 1A and 1C). None of the infants had SARS-CoV-2-specific IgA in the blood, but surprisingly, half of the infants in the Antepartum group had high titer SARS-CoV-2-specific IgA in the nose that exceeded titers found in breastmilk (Figure 1B and 1D). None of the infants in the Postpartum group had SARS-CoV-2-specific IgA in the nose (1B and 1D).

**Table 1.** Demographic information

|  | Antepartum vaccination<br>n=10 | Postpartum vaccination<br>n=12 |
| --- | --- | --- |
| Mother age, median (range) | 33 (27-37) | 34 (29-40) |
| Mother race |  |  |
| White, n (%) | 9 (90) | 12 (100) |
| Black, n (%) | 1 (10) | 0 (0) |
| Asian, n (%) | 0 (0) | 0 (0) |
| Mother, history of COVID-19, n (%) | 0 (0) | 0 (0) |
| Vaccine type |  |  |
| Pfizer, n (%) | 5 (50) | 5 (42) |
| Moderna, n (%) | 5 (50) | 7 (58) |
| Vaccine booster (postpartum) |  |  |
| Pfizer, n (%) | 2 (20) | 0 (0) |
| Moderna, n (%) | 1 (10) | 0 (0) |
| Infant gestational age at birth, median (range) | 39 (36-41) | 39 (37-40) |
| Infant race |  |  |
| White, n (%) | 8 (80) | 11 (92) |
| Black, n (%) | 2 (20) | 0 (0) |
| Asian, n (%) | 0 (0) | 1 (8) |
| Infant sex |  |  |
| Male, n (%) | 5 (50) | 5 (42) |
| Female, n (%) | 5 (50) | 7 (58) |
| Infant, history of COVID-19, n (%) | 0 (0) | 0 (0) |
| Infant sampling age | 125 (15 to 217) | 350 (209 to 545) |
| Timing of vaccine to birth in days, median (range) | -175 (-42 to -285) | 170 (26 to 326) |
| Timing of vaccine to infant sampling in days, median (range) | 287 (98 to 335)* | 201 (167 to 232)** |
| Number of ounces of breastmilk intake/day, median (range) | 27 (0-30)*** | 25 (0-32)**** |

\* Mother plasma and milk sampling done at same time as infant.
\*\* Mother milk sampling done at same time as infant, but blood sampling done at median of 125 days from vaccine (60 to 137)
\*\*\* One of the infants in the antepartum vaccinated stopped breastfeeding several weeks prior to sampling.
\*\*\*\* Three Infants in the postpartum vaccinated groups discontinued breastfeeding 2-3 months prior to sampling.

**Figure 1.**
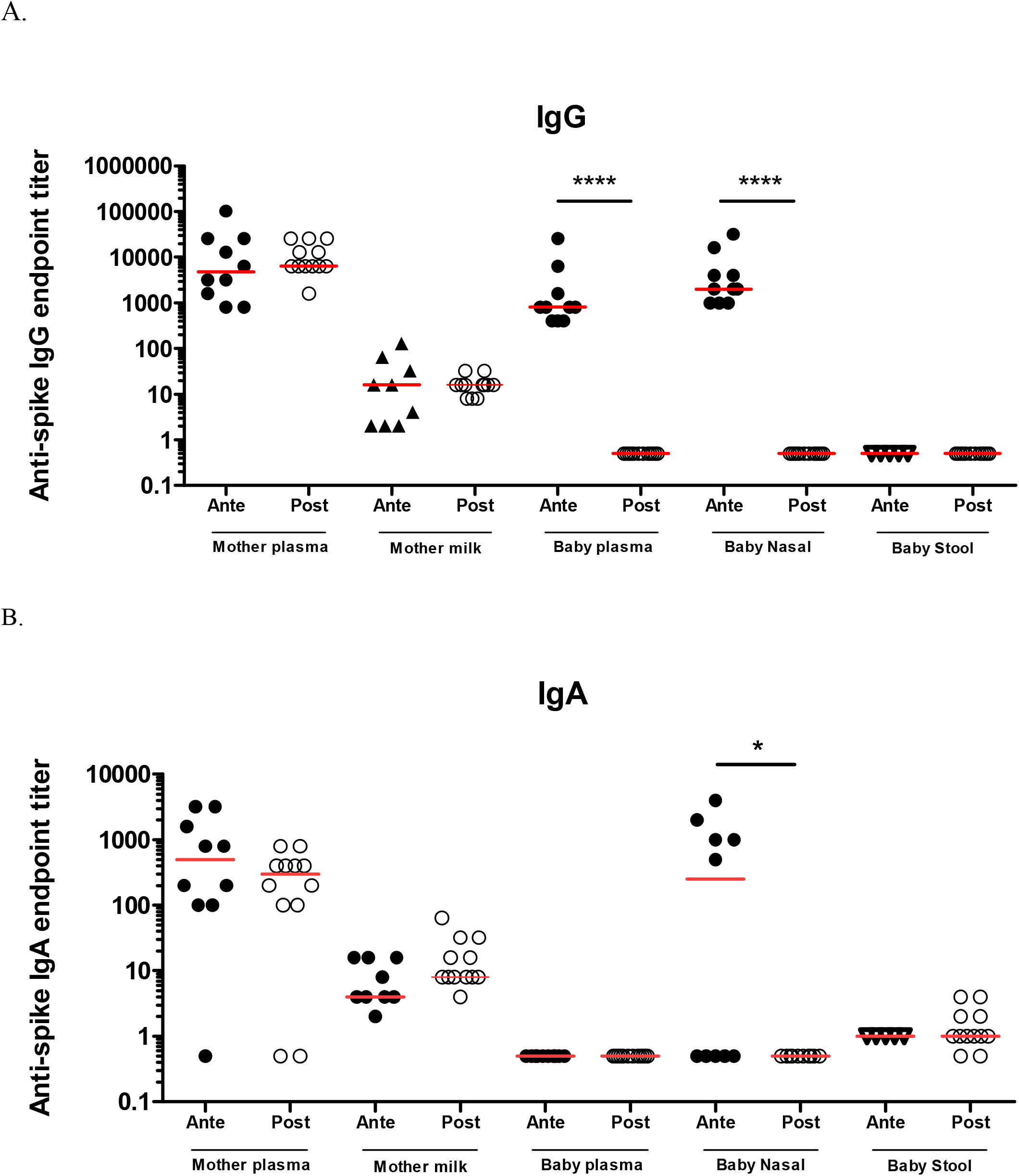
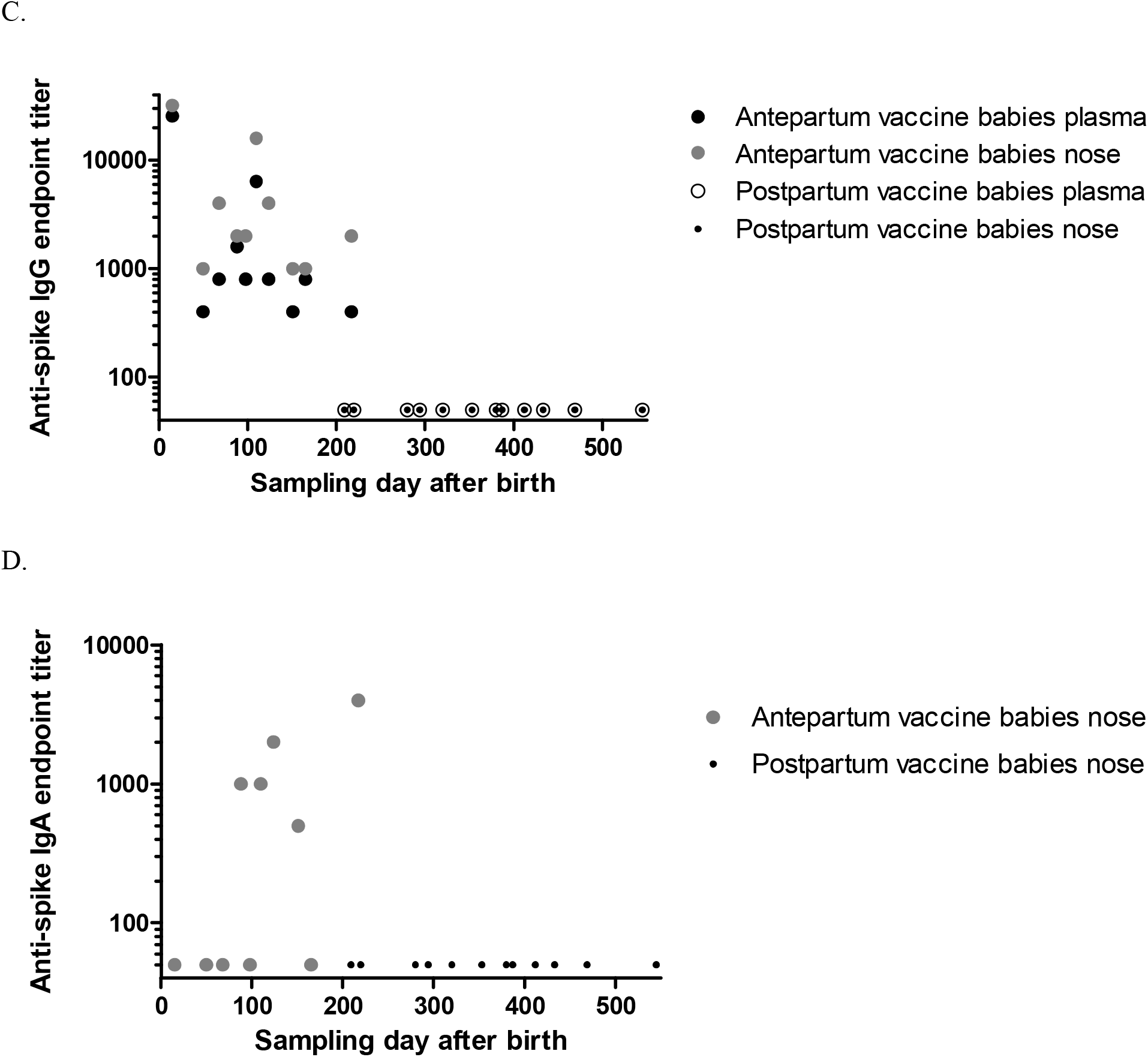
SARS-CoV-2 spike binding titers in vaccinated mothers and infants. A) Log IgG binding titer in mother plasma, mother milk, infant plasma, infant nose, and infant stool. Samples below the limited of detection given an artificial value of 0.5. Horizontal red lines represent median values. B) Log IgA binding titer in mother plasma, mother milk, infant plasma, infant nose, and infant stool. Samples below the limited of detection given an artificial value of 0.5. Horizontal red lines represent median values. C) Infant plasma and nasal IgG log transformed and plotted (Y-axis) vs sampling day (X-axis). Samples below the limited of detection given an artificial value of 45. D) Infant nasal IgA log transformed and plotted (Y-axis) vs sampling day (X-axis). Samples testing negative given an artificial value of 50. All Elisas modified from an ELISA designed to detect SARS-CoV-2 spike trimer as previously described, samples run in duplicate, and all results reported as endpoint titers. Limit of detection of the IgG plasma, milk, nasal, and stool assays were 1:50, 1:1, 1:1000, and 1:1, respectively. Limit of detection of the IgA plasma, milk, nasal, and stool assays were 1:100, 1:1, 1:500, and 1:1, respectively. Differences between groups were tested by the 2-tailed Mann-Whitney test, with a P ≤ .05 being considered significant. * = P ≤ 0.05; ** = P ≤ 0.01; *** = P ≤ 0.001; **** = P ≤ 0.0001

## Discussion

These data suggest that antepartum vaccination followed by breastfeeding provides the most reliable source of systemic and mucosal antibodies against SARS-CoV-2 for infants. Breastfed infants only acquired systemic anti-SARS-CoV-2 antibodies if their mothers were vaccinated antepartum. Several studies have shown systemic absorption of antibodies through the gastrointestinal tract in preterm newborns, which ceases after the first day of life in term newborns.^4^ However, breastfeeding in and of itself provides protection against certain upper respiratory infections,^5^ through an unknown mechanism. In this study, older infants (all of whom were postpartum vaccination group) had no detectable nasal antibodies to SARS-CoV-2. However, we detected vaccine-specific IgA at high concentrations in nasal samples in about half the younger infants whose mothers were vaccinated antepartum and breastfed. This is noteworthy because none of the infants had detectable systemic, vaccine-specific IgA and thus had not obtained it transplacentally but rather by breastfeeding. Moreover, their nasal, vaccine-specific IgA titers were higher than corresponding breast milk titers. Pharyngonasal reflux or direct inhalation could directly deposit the vaccine-specific IgA (and IgG) on nasal surfaces, but another mechanism for concentrating the antibodies (such as binding to mucin or lectin) would need to be involved. Alternatively, plasmablasts from the mother could be transferred with the colostrum via breastfeeding immediately postpartum and the vaccine-specific IgA produced locally.^4,6^

Limitations of this study include difference in ages of the infants in the two groups (partly due to vaccine schedule), cross-sectional design, small study population, and small sample volume that precluded functional testing. While breastfeeding brings many benefits for the infant, there does not appear to be any meaningful transfer of SARS-CoV-2 specific antibody from mothers to their infants if vaccinated postpartum. In mothers who breastfed, vaccination antepartum followed by breastfeeding appears to be the best way to provide systemic and local anti SARS-CoV-2 antibodies for infants.

## Data Availability

All data produced in the present study are available upon reasonable request to the authors

## Acknowledgements

We would like to thank all of the study participants who donated their time and samples. We would like to also thank Zahra Rikhtegaran Tehrani, PhD; Madeleine Purcell, BS; James D. Campbell, MD; Kristopher B. Deatrick, MD; Casey Bor, CRNP; Dawn M. Parsell, MS, PA-C; Colleen Dugan MS, AG-ACNP; Andrea R. Levine, MD; Sabrina C. Ramelli, PhD; Daniel S. Chertow, PhD; and Daniel L Herr, MD, who helped with study logistics. This work was supported in part by Merit Award # I01 BX005469-01 from the United States (U.S.) Department of Veterans Affairs Biomedical Laboratory Research and Development Service. The contents do not represent the views of the U.S. Department of Veterans Affairs or the United States Government.

## References

1. American College of Obstetricians and Gynecologists. Practice advisory: COVID-19 vaccination considerations for obstetric–gynecologic care. In:2020.

2. Saadat S, Rikhtegaran Tehrani Z, Logue J, et al. Binding and Neutralization Antibody Titers After a Single Vaccine Dose in Health Care Workers Previously Infected With SARS-CoV-2. JAMA. 2021;325(14):1467–1469.

3. Rikhtegaran Tehrani Z, Saadat S, Saleh E, et al. Performance of nucleocapsid and spike-based SARS-CoV-2 serologic assays. PLoS One. 2020;15(11):e0237828.

4. Ogra SS, Weintraub D, Ogra PL. Immunologic aspects of human colostrum and milk. III. Fate and absorption of cellular and soluble components in the gastrointestinal tract of the newborn. J Immunol. 1977;119(1):245–248.

5. Downham MA, Scott R, Sims DG, Webb JK, Gardner PS. Breast-feeding protects against respiratory syncytial virus infections. Br Med J. 1976;2(6030):274–276.

6. Atyeo C, Alter G. The multifaceted roles of breast milk antibodies. Cell. 2021;184(6):1486–1499.

